# Learning Patient-Specific Event Sequence Representations for Clinical Process Analysis

**DOI:** 10.64898/2026.03.25.26348333

**Authors:** Katalin Sólyomvári, Tuomo Antikainen, Hans Moen, Pekka Marttinen, Risto Renkonen, Miika Koskinen

## Abstract

Healthcare system performance evaluation is constrained by episodic performance indicators and process mining techniques that fail to accommodate the scale, heterogeneity, and temporal complexity of real-world clinical pathways. Electronic health records enable reconstructing patient journeys that capture how care processes unfold across fragmented healthcare services. Here we present ClinicalTAAT, a time-aware transformer that bridges clinical sequence modeling and process mining by integrating contextual and time-varying information to learn interpretable patient-specific representations from inherently sparse, irregular and high-dimensional clinical event sequences. Evaluated on a large pediatric emergency cohort, ClinicalTAAT outperforms existing models in acuity and diagnosis classification, identifies clinically meaningful patient subgroups in heterogenous population with distinct acuity, resource utilization and diagnostic patterns, and detects anomalies in individual care trajectories. These findings demonstrate that time-aware transformers can complement existing process mining methodologies and serve as foundation models for clinical process analysis, providing a scalable framework for data-driven healthcare evaluation and optimization.

## 1 Introduction

Healthcare systems face rising costs and growing demand for services while striving to deliver individualized high-quality care. The fragmented healthcare system consists of interdependent subprocesses whose localized policies and collective performance shape delays, bottlenecks, and patient outcomes across the entire care journey. The fragmented healthcare system involves interconnected subprocesses, whose cooperative efficiency and local policies are cumulatively reflected in experienced delays, bottlenecks, and patient outcomes throughout their journeys. Designing efficient and seamless individual-level service processes is further complicated by population heterogeneity and varying service needs. Consequently, traditional performance indicators are predominantly structured around isolated point-in-time metrics that offer limited capacity for longitudinal monitoring and fail to reflect broader patterns of resource utilization and system-level inefficiency [1].

Effective system-level management therefore requires longitudinal representations of patient journeys that account for cross-sector service use, care sequencing, and cumulative exposure in a heterogeneous population. While Electronic Health Records (EHRs) provide extensive real-world clinical data, patient trajectory modeling remains technically demanding due to the stochasticity, temporal sparsity, and high dimensionality of multi-departmental event sequences. Thus, there is a critical need for advanced computational frameworks capable of capturing the real-world complexity of patient journeys and transforming this information into practical insights for systematic evaluation and optimization of service processes.

Contemporary process mining and machine learning frameworks for process modeling address only a subset of the challenges inherent in irregular, heterogeneous clinical event data. Process mining facilitates the extraction of structured representations from event logs, supporting the discovery, monitoring, and enhancement of clinical workflows [2–6]. Within the healthcare domain, these methodologies have effectively characterized care pathways, identified operational bottlenecks, and evaluated institutional workflow conformance at the population level [2, 7, 8]. By abstracting event sequences into aggregated workflow models that capture organizational structures and normative care patterns across patient cohorts [9], process mining provides valuable insights into institutional care delivery. However, this aggregation-centric paradigm presents significant limitations in highly heterogeneous clinical settings characterized by complex longitudinal patient pathways. To maintain interpretability, current process mining models frequently constrain the analysis to specific subtasks or employ substantial abstraction, event filtering, and trace clustering [10]. Such techniques may obscure individual patient trajectories, compromising patient-level granularity and temporal precision [2]. Consequently, conventional process mining approaches fall short in representing prolonged and highly variable patient journeys across fragmented healthcare systems and do not provide sufficient support for individual-level predictive modeling [7].

In contrast, deep learning architectures, particularly Transformer models, have shown promise in capturing intricate sequential dependencies and population heterogeneity. By learning vector representations of patient trajectories, these models enable sophisticated embedding-based analyses. However, despite their representational power, Transformer frameworks face significant challenges when applied to complex healthcare processes, such as managing highly irregular timing and the large variety of clinical event types characteristic of real-world medical data.

Despite recent advances, clinical transformer-based modeling faces four major methodological gaps. First, standard transformer architectures typically assume ordinal token relationships rather than modeling actual temporal intervals, or they use simplified temporal encodings such as discrete time bins. Second, although representation learning holds promise for patient stratification, transformers have rarely been leveraged to gain insights into sequential patterns or clustering [11]. Existing literature tends to prioritize downstream task performance, often resulting in a lack of systematic evaluations regarding the quality and clinical relevance of learned representations [12–15]. Third, the interpretability of these models frequently relies on post-hoc visualization of attention mechanisms [16] rather than rigorous analytical frameworks designed to extract structured care patterns. Fourth, while techniques for integrating static and temporal variables have been proposed [17–19], there remains a lack of structured fusion architectures that explicitly characterize the complex interactions between these data modalities. As a result, these models often fail to capture the irregular and clinically significant temporal dynamics inherent to longitudinal patient trajectories [20, 21]. Although recent time-aware transformer variants have emerged, their focus remains predominantly on continuous-valued forecasting rather than on synthesizing interpretable representations for categorical event sequences in clinical pathway analysis [22–26].

In this study, we propose ClinicalTAAT (C-TAAT), a bidirectional representation learning framework for clinical event sequences based on a Time-Aware Attention-based Transformer (TAAT)[27]. C-TAAT explicitly incorporates irregular inter-event intervals through specialized time-aware attention mechanisms and leverages cross-attention to integrate static patient-level covariates. This architecture enables the mapping of diverse longitudinal patient journeys within a population and creates latent representations that capture time-related details more accurately than conventional transformer models. Our primary objective is to evaluate the efficacy of this framework in learning interpretable care sequence representations that facilitate subgroup discovery, with an emphasis on structural insights rather than solely optimizing individual task-specific performance. Our evaluation is structured around four foundational research questions:

1. Do the learned embeddings capture clinically meaningful structure from pediatric emergency sequences, as revealed by cluster analysis?
2. How effectively do pretrained representations transfer to downstream tasks such as ESI acuity and diagnosis classification via fine-tuning?
3. What clinical patterns and temporal dynamics drive model predictions, as analyzed through feature attribution and predictability?
4. Do the representations align with known clinical pathways when evaluated on synthetic data with controlled temporal structures?

By bridging patient-level representation learning with process-oriented analysis of clinical care pathways, this work provides a foundation for data-driven evaluation and optimization of healthcare delivery, with direct relevance for understanding variation and inefficiency in real-world clinical processes.

## 2 Methods

### ClinicalTAAT architecture

We introduce ClinicalTAAT, a clinical adaptation of the Time-Aware Attention-based Transformer (TAAT) [27] designed for modeling emergency care event sequences (Figure 1).

**Figure 1:**
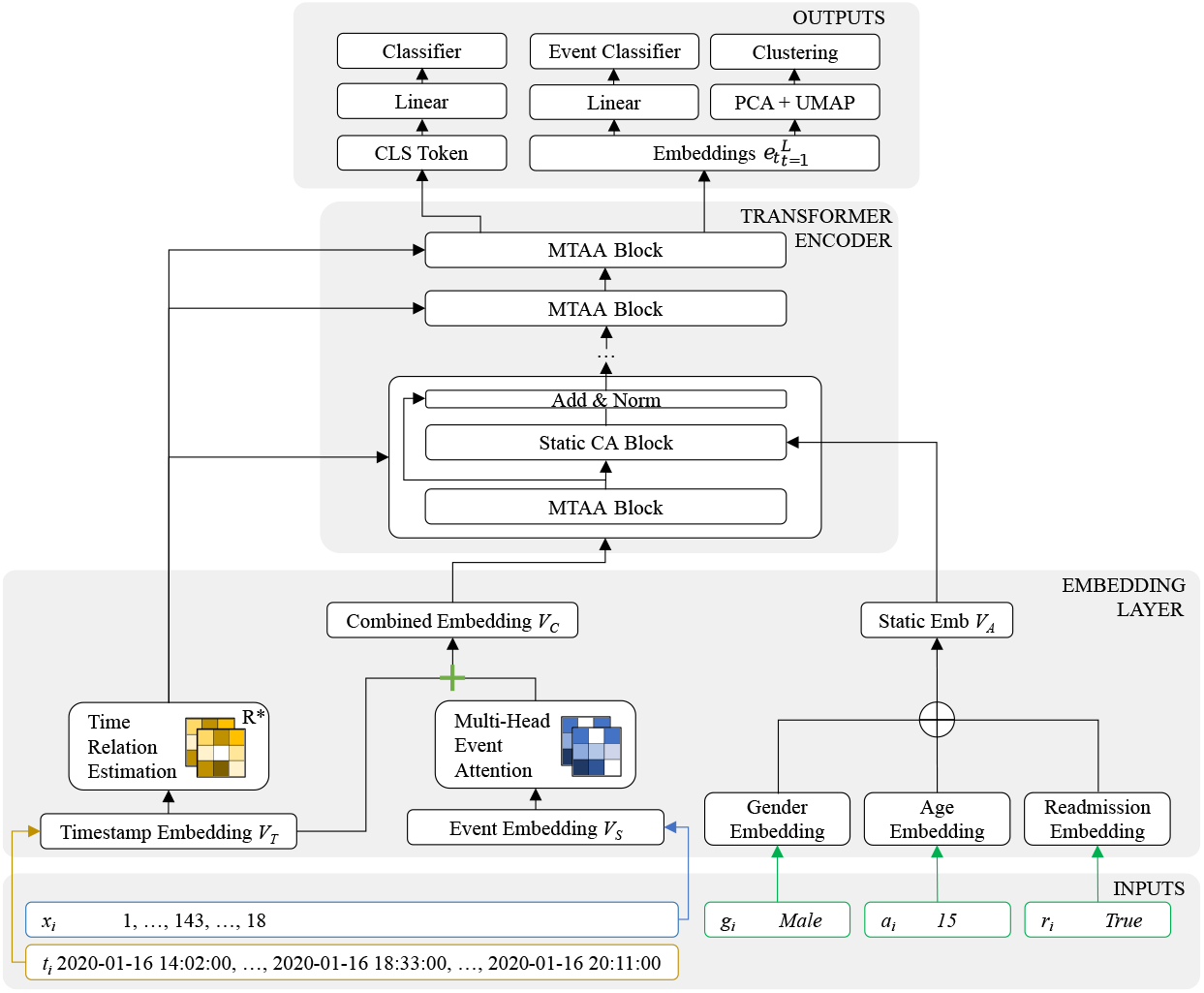
Model architecture. Multi-head Time-Aware Attention (MTAA) blocks process timestamped clinical event embeddings. Static features are integrated via cross-attention (CA).

#### Input representation and modeling approach

For each patient *p* in a cohort, the model takes as input a sequence of timestamped clinical events 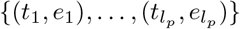 along with static contextual features **s**_*p*_, where the sequence length *l*_*p*_ varies across patients. Following process mining conventions for activity logs, the analysis was restricted to (time, event) tuples without including the value component [28]. To create a fixed-length input for the transformer, a special classification token `[CLS]` was prepended and padding was added with `[PAD]` tokens in the end, resulting in the processed sequence: 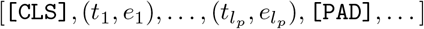. The model employs TAAT’s multi-granularity temporal encoding [27], which decomposes relative time differences Δ*t* into *ϕ* discrete components (days, hours, minutes, seconds):

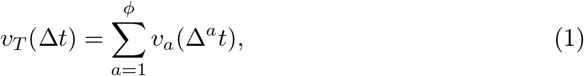

where *v*_*a*_(·) is a learnable embedding function for the *a*-th temporal component. This approach handles irregular clinical timing patterns inherent to emergency care sequences, where timing reflects critical workflow dependencies.

#### Time-aware attention

Time-aware attention [27] integrates temporal relationships directly into the self-attention calculation:

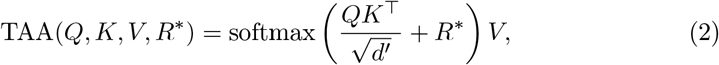

where *Q, K*, and *V* are learnable query, key, and value matrices computed from event embeddings via linear projections, and *R*^*^ ∈ ℝ^*L*×*L*^ is a temporal relation matrix computed via the Time Relation Estimation (TRE) module:

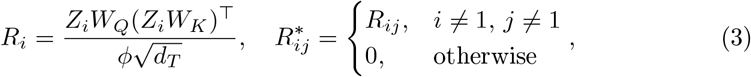

with 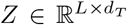 the embedded timestamp sequence, 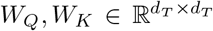 learnable projection matrices, and the zeroing operation ensuring the class token does not contribute to temporal relations.

#### Static feature integration

A cross-attention mechanism is employed to integrate static patient features, allowing the model to condition temporal processing on patient-specific context. Static features (age, gender, readmission indicator) are embedded into vector *V*_*A*_ and integrated:

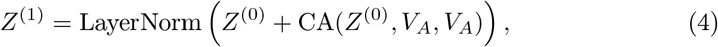

where 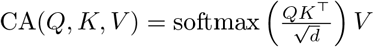 with *Q* as the event sequence embedding and *K, V* as projections of *V*_*A*_, the broadcasted static feature embedding. This approach allows the model to dynamically adjust temporal attention based on static patient context.

### Model training

Our training follows a representation learning paradigm with two distinct phases. (i) Following BERT’s strategy [29], during self-supervised pretraining the model learns through masked event prediction in which 15% of tokens were masked, of which 80% were replaced with `[MASK]`, 10% with a random token, and 10% remained unchanged. (ii) At the supervised finetuning phase, the pretrained model is adapted for down-stream prediction. The `[CLS]` token, is linked to separate classification heads: one for ESI triage and another for diagnosis category prediction.

To address class imbalance, we employed focal loss [30] for both pretraining and supervised tasks. Detailed training and optimization settings are provided in Appendix Section A.3.

In order to compare the performance of our fine-tuned models, we adapted two established clinical transformer architectures, BEHRT [20] and STraTS [25], to serve as baselines on both ESI acuity and diagnosis classification tasks. For fair comparison, both models were modified to accept timestamped event sequences by processing (time, event) tuples. Implementation details, including temporal embedding adaptations and static feature integration, are provided in Appendix Section A.5.

### Patient representation clustering workflow

To assess whether self-supervised pretraining yields clinically meaningful representations, we applied clustering analysis to the learned embeddings. Our aim was to determine whether the model intrinsically groups patients according to clinical acuity, diagnostic trends, and resource usage: dimensions never provided during training.

We extracted latent representations **e**_*t*_ ∈ ℝ^*d*^ (with *d* = 64) for each event from the final encoder layer and concatenated them across the sequence, yielding a patient-level embedding **p**_*i*_ ∈ ℝ^*Ld*^ where *L* = 257 is the maximum sequence length. This produced a patient embedding matrix **P** ∈ ℝ^*N* ×*Ld*^ for the full cohort of *N* = 227, 782 patients. Following typical workflow [31], we first applied principal component analysis (PCA) to reduce dimensionality to 50 components (preserving 96% variance), then further to reduce to 2 dimensions using UMAP to preserve local and global topological structure. Finally, we applied BIRCH clustering [32] to identify subgroups of patient trajectories in the reduced space.

We quantitatively evaluated cluster quality and clinical coherence using established clustering metrics and baseline comparisons, detailed in Section 3. This pipeline also enabled comparison between learned embeddings and traditional feature engineering approaches (e.g aggregated counts and summary statistics), revealing whether self-supervision captures clinically relevant care patterns absent from manually constructed features (see Appendix Section A.5.1 for details).

### Interpretability analyses methods

To understand the contributions of single events on predictions, we performed Shapley Additive exPlanations (SHAP) analysis [33] on the classifiers fine-tuned for ESI acuity and diagnosis classification using `KernelExplainer`, which quantifies each event’s marginal contribution to the model’s confidence via weighted linear regression on perturbed input samples relative to a baseline of typical patient sequences. For a patient’s event sequence *S* = [*e*_1_, *e*_2_, …, *e*_*n*_], let *f* (*S*) denote the model’s maximum softmax probability for its predicted class. The SHAP value *ϕ*_*i*_ for event *e*_*i*_ quantifies how much *e*_*i*_ increases (*ϕ*_*i*_ > 0) or decreases (*ϕ*_*i*_ < 0) the model’s confidence relative to a baseline expectation. This baseline is the average model prediction over a background dataset representing typical patient trajectories. We constructed this background by sampling 100 patient sequences and adjusting them to a common length, ensuring clinical plausibility.

To quantify the sensitivity of the learned representations to different types of sequence modifications, we measured embedding displacements under controlled perturbations (e.g., event changes, timing shifts) using Euclidean distance in the reduced UMAP space, the same metric used for silhouette score computation, providing consistency with our clustering evaluation.

To test whether C-TAAT can detect uncommon sequences that deviate from standard pathways and to identify implausible individual events or timing, we measured the predictability of individual events under systematic perturbations. For each event in a patient’s sequence, we masked it and computed the model’s predicted probability for the true event, providing a quantitative measure of pathway plausibility.

### Datasets

We evaluated model performance using a real-world dataset comprising pediatric emergency care encounters at HUS Helsinki University Hospital, Finland, collected between February 2020 and February 2024. For each hospitalization episode, we extracted the full temporal sequence of clinical events until the discharge from hospital, together with associated outcomes. The event vocabulary consisted of |*V*| = 337 distinct clinical activities, including laboratory tests, imaging examinations, bedside procedures, and physician encounters. Time-fixed covariates included age (0–16 years), sex, and 30-day readmission status. To ensure computational tractability, we retained encounters with 3–256 recorded events, covering 99.76% of all visits. Timestamps were normalized to minutes elapsed since admission. Event sequences were padded to a uniform length of 256 and partitioned into training (70%), validation (15%), and test (15%) subsets. Figure 2 presents an overview of the preprocessing pipeline for a representative patient trajectory. The dataset supported three tasks: (i) masked event prediction during pretraining and (ii) Emergency Severity Index (ESI) triage classification and (iii) diagnosis category prediction during fine-tuning.

**Figure 2:**
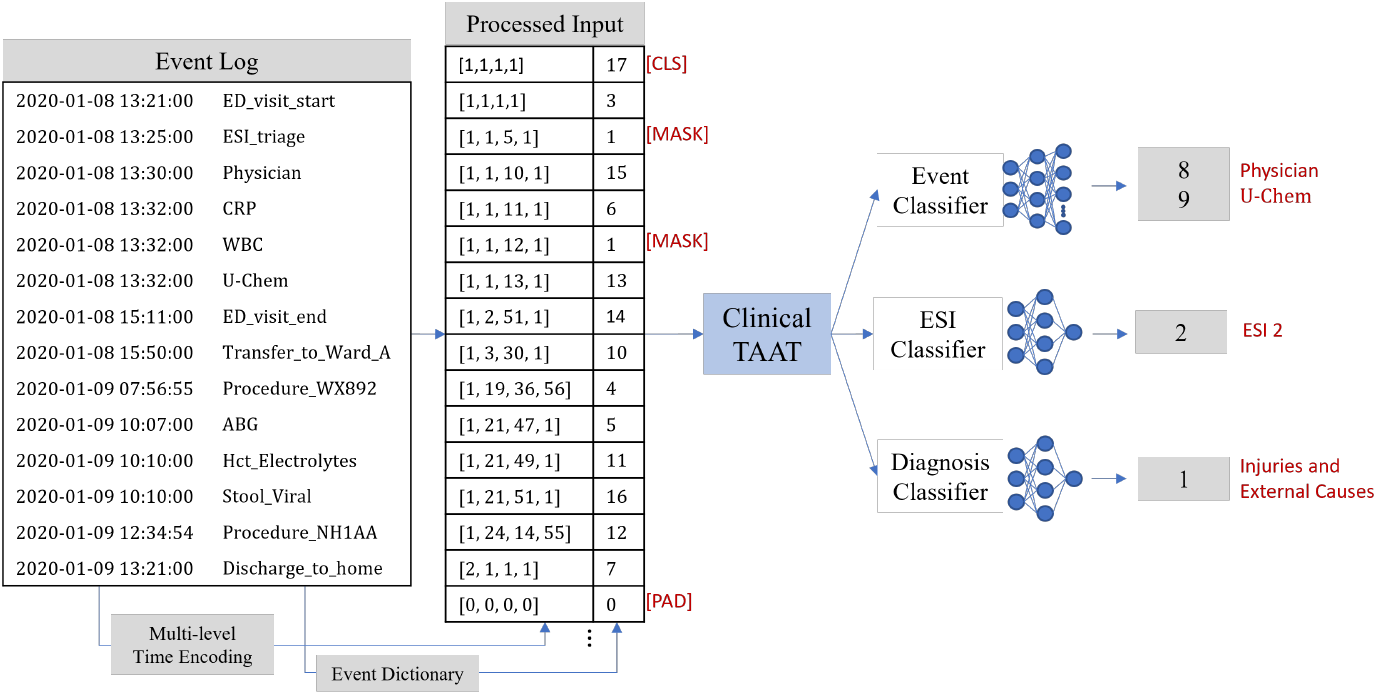
Data preprocessing pipeline (representative synthetic encounter). Timing of events (left) were tokenized and combined with relative timing information, created via C-TAAT’s multi-level time-encoding (center). Processed sequences served as input to ClinicalTAAT (right) for masked event pretraining and fine-tuned ESI or diagnosis classification.

In addition, we constructed a controlled synthetic emergency care dataset with established ground-truth temporal patterns, designed to verify that the model can learn clinically relevant temporal dependencies and anomalies in patient journeys. This synthetic data enabled systematic validation under controlled conditions, testing whether learned representations recover known care patterns, temporal dynamics, and pathway structure regardless of their specific clinical accuracy. We generated sequences with simplified but structured patterns using a care pattern- and diagnosis-conditioned second-order Markov process, producing distinct and interpretable care pathways across different simulated trajectory types. Each sequence was annotated with clinically relevant labels, including Emergency Severity Index (ESI), diagnosis category, and 30-day readmission risk, along with static patient attributes (age and gender).

## 3 Results

### Dataset characteristics

The real-world dataset comprised 227,782 pediatric emergency encounters of of 111,061 patients, with sequence lengths varying considerably across patients (3–256 events per encounter) and irregular temporal intervals between events. The dataset included two complementary clinical labels for each patient encounter: the 5-level Emergency Severity Index (ESI) triage acuity score (1: most urgent to 5: non-urgent), with a distribution of ESI 1: 0.5%, ESI 2: 7.1%, ESI 3: 26.3%, ESI 4: 33.2%, and ESI 5: 33%; and 13 primary diagnosis categories based on ICD-10 chapters. The most frequent diagnostic categories were Cardiovascular and pulmonary diseases (ICD chapters I and J), Injuries and external causes (ICD chapters S to Y), Special codes (ICD chapters U and Z), Other symptoms and signs (ICD chapter R), and Sensory disorders (ICD chapter H). Appendix Table A2 provides a detailed breakdown of diagnosis categories.

We generated 230,000 synthetic patient sequences. Temporal dynamics explicitly varied by trajectory type, capturing regular, irregular, and burst-driven patterns that reflect differences in care intensity. Shorter sequences were generated to ensure pattern clarity and minimize noise, focusing validation on systematic structure recovery rather than sequence length extremes. Summary statistics with full generation details are provided in Appendix Section A.1.

### Self-supervised learning of sequence patterns

Pretraining performance was assessed using masked event prediction as a classification task on real-world clinical data (Table 1). The model achieved 88% accuracy in predicting masked clinical events with a top-5 accuracy of 97%, including infrequent events. Thus, the learned representations effectively captured contextual patterns of event occurrence throughout the entire event vocabulary. The model demonstrated robust performance across multiple event types, as evidenced by a weighted precision and recall score of 0.86, adjusted for class frequency. The lower macro F1-score, which weights each class with equal importance, compared to weighted metrics reflects the imbalanced nature of clinical event distributions in our dataset. The strong weighted F1-score indicates that the model effectively captured general patterns in frequently occurring clinical events, while lower recall for rare events reflects the highly imbalanced distribution across more than 300 event types. Together, these results demonstrate that C-TAAT learns meaningful sequence patterns suitable for embedding analysis and downstream tasks, a finding further supported by the ablation study in Appendix Section A.6.2.

**Table 1:**
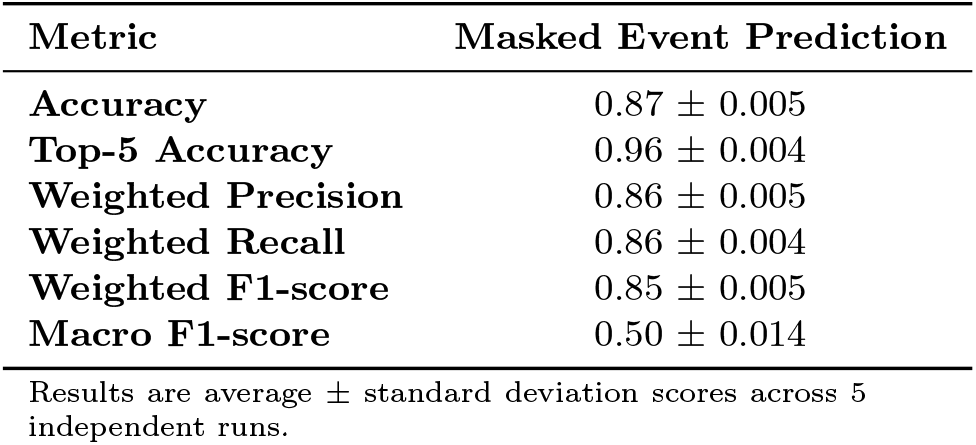
Self-supervised pretraining performance on masked event prediction.

| Metric | Masked Event Prediction |
| --- | --- |
| **Accuracy** | $0.87 \pm 0.005$ |
| **Top-5 Accuracy** | $0.96 \pm 0.004$ |
| **Weighted Precision** | $0.86 \pm 0.005$ |
| **Weighted Recall** | $0.86 \pm 0.004$ |
| **Weighted F1-score** | $0.85 \pm 0.005$ |
| **Macro F1-score** | $0.50 \pm 0.014$ |

Results are average ± standard deviation scores across 5 independent runs.

### Heterogeneity of patient trajectories

Birch clustering of representation vectors learned through self-supervised pretraining identified 17 clinically meaningful patient subgroups (Figure 3). These vectors, which serve as concise summaries of sparsely sampled clinical sequences, corresponded to distinct care patterns. The clusters exhibited systematic differences in clinical acuity and diagnoses (Figure 4a-b). High-acuity clusters, such as clusters 1 and 16, contained a higher proportion of ESI 1–2 patients, while low-acuity clusters, such as clusters 7 and 14, comprised primarily ESI 4–5 cases. Resource utilization followed the same pattern: high-acuity clusters showed elevated use of imaging, lab tests, and consultations, whereas low-acuity clusters showed minimal consumption of resources (Figure 4c). Quantitative evaluation confirmed that the clusters were well-separated and internally coherent, with silhouette and Davies–Bouldin scores outperforming baseline methods (Appendix Table A5).

**Figure 3:**
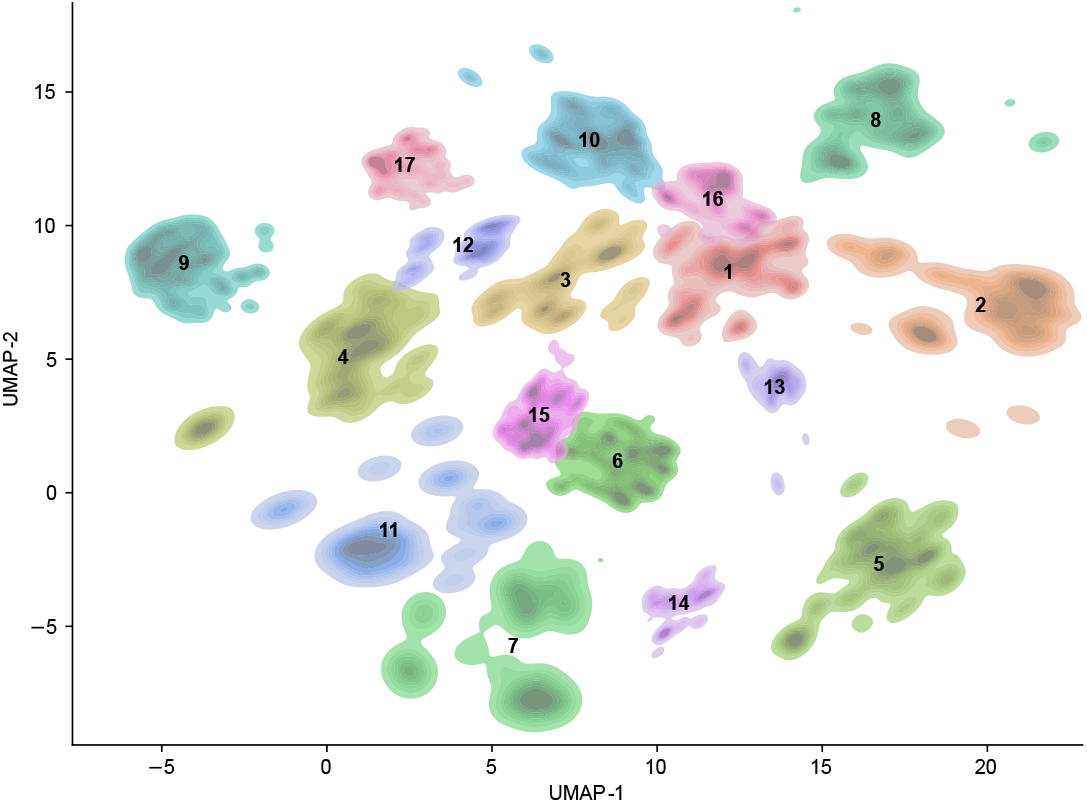
Clustering of patient-specific representation vectors reveals distinct care pattern subgroups.

**Figure 4:**
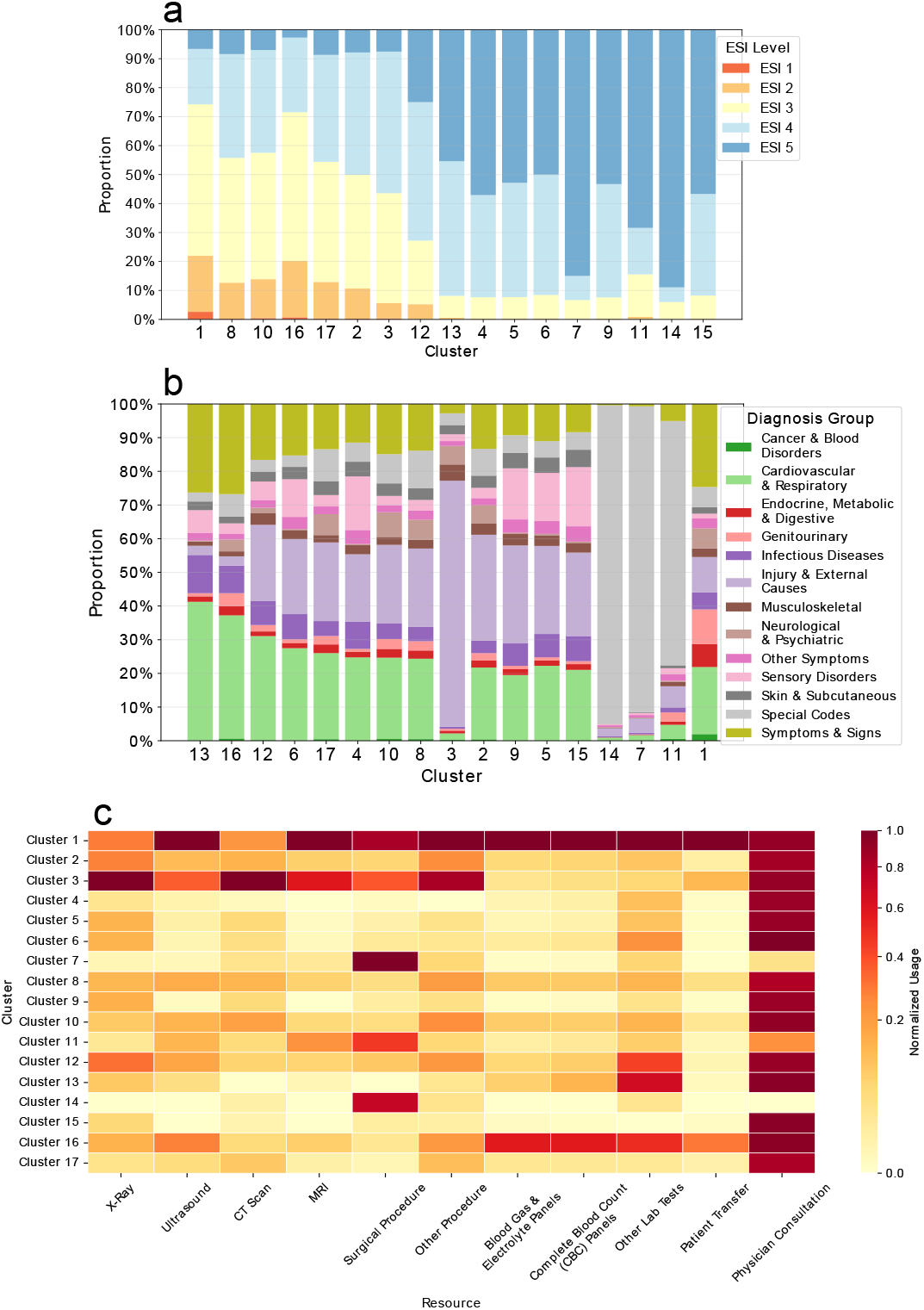
Clustering analysis of pretrained C-TAAT trajectory embeddings. (a) ESI score distribution per cluster. (b) Diagnosis category distribution per cluster. (c) Heatmap of normalized resource utilization across clusters, scaled column-wise to [0, 1] relative to the maximum utilization observed for each resource. Clusters represent distinct care patterns with clear acuity stratification, clinical presentation patterns, and associated resource utilization.

Clusters containing high-acuity patients were also associated with elevated readmission rates (21–23% vs. 16% average), longer care sequences, and distinct demographic profiles. Cluster 1 comprised older children with diverse acute presentations, while Cluster 16 featured younger children with respiratory infections. Specific injury patterns also emerged; for example, Cluster 3 captured older pediatric patients with upper extremity orthopedic trauma. Conversely, clusters 7, 11, and 14 represented low-complexity cases characterized by counseling and administrative encounters (Z71 diagnoses), minimal resource use, and the lowest readmission rates (11–12%). These care patterns demonstrate that the embeddings capture the full spectrum of emergency care—from acute interventions to preventive visits—without explicit supervision for these distinctions.

A detailed summary of all cluster characteristics is provided in Appendix Table A8.

### Classification performance

After finetuning for classification tasks, C-TAAT achieved 62% accuracy on ESI acuity classification and 49% accuracy on diagnosis classification. Comparing our model against adapted versions of BEHRT and STraTS on real-world data (Table 2) revealed that for ESI classification, a task intrinsically tied to event timing, C-TAAT achieved considerable improvements over both baselines across all metrics. For diagnosis classification, C-TAAT outperformed STraTS, but showed only minimal advantage over BEHRT.

**Table 2:** Comparison against clinical transformer baselines.

| Task | Model | Accuracy | Macro F1 | Weighted F1 |
| --- | --- | --- | --- | --- |
| ESI | **ClinicalTAAT** | **0.62 ± 0.000** | **0.57 ± 0.002** | **0.61 ± 0.001** |
| | Adapted-BEHRT | 0.60 ± 0.001 | 0.53 ± 0.007 | 0.59 ± 0.001 |
| | Adapted-STraTS | 0.59 ± 0.003 | 0.50 ± 0.016 | 0.57 ± 0.003 |
| Diagnosis | **ClinicalTAAT** | **0.49 ± 0.007** | **0.38 ± 0.006** | **0.49 ± 0.004** |
| | Adapted-BEHRT | 0.48 ± 0.002 | 0.38 ± 0.004 | 0.48 ± 0.001 |
| | Adapted-STraTS | 0.47 ± 0.002 | 0.37 ± 0.005 | 0.47 ± 0.004 |

Values show mean ± s.d. across 3 runs.

### Interpretability on synthetic data

To validate model behavior against known ground truth while eliminating real-world confounders, we conducted controlled experiments on synthetic data with encoded trajectory types, testing whether the model recovers these predefined patterns. Clustering of learned sequence vectors revealed patterns consistent with our real-world findings (Appendix Figure A5), confirming that the model captures clinically meaningful structure in a controlled setting.

#### Single event contributions to predictions

SHAP analysis on the fine-tuned classifiers revealed the clinically relevant events for each prediction task. Diagnosis predictions relied on diagnosis-specific events (like cardiac medications for cardiac patients), while ESI acuity predictions emphasized critical events and interventions such as intravenous access and airway management. Events unrelated to the patient’s primary diagnosis (e.g., routine labs in critical cases) showed minimal influence on predictions (see Appendix Section A.8 for detailed examples).

#### Influence of event timings on representations

To test whether timing adds predictive value beyond event occurrence alone, we compared our model against a version pretrained without temporal encoding on synthetic data, evaluating performance on masked event prediction. The temporal model improved top-1 accuracy by 13.3% (0.627 vs. 0.494) and weighted F1 by 14.5% (0.619 vs. 0.474), confirming that temporal relationships are critical for learning event sequences.

#### Embedding sensitivity to perturbations

To determine whether learned representations reflect both clinical content and temporal structure, and which dominates, we systematically perturbed synthetic patient sequences. Embedding sensitivity to perturbations varied across modification types (Figure 5): event changes produced the largest embedding displacements, while other perturbations—including random timing, interval increases, and last event shifts—resulted in smaller but measurable effects. Full perturbation descriptions and quantitative results are provided in Appendix Section A.8.

**Figure 5:**
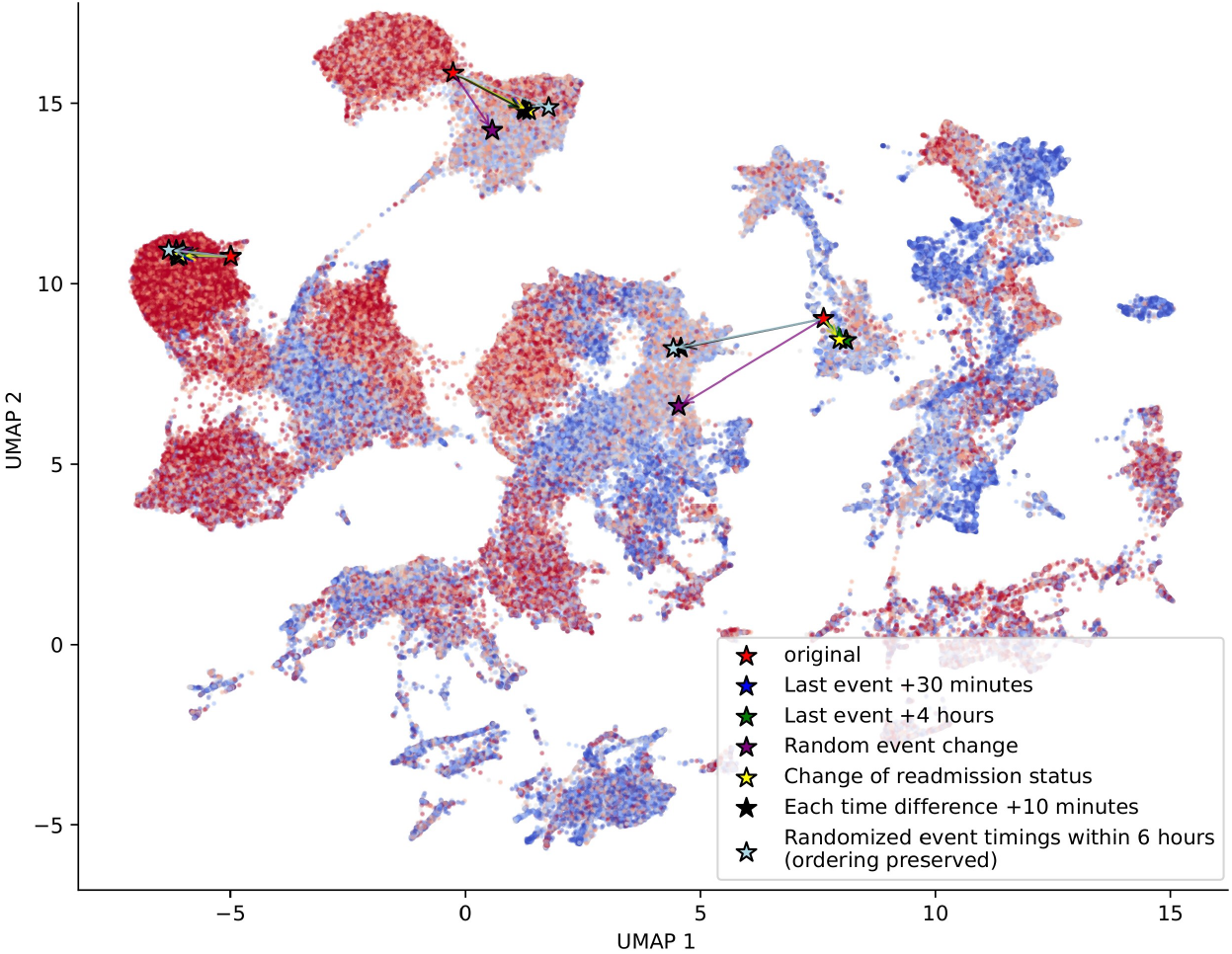
Sensitivity analysis of embeddings under systematic perturbations. Event changes caused the largest displacements, followed by substantial temporal shifts (e.g., +6h, randomized timing). Minor timing variations (e.g., +60min) produced negligible effects, demonstrating understanding of clinical content and temporal patterns.

#### Anomaly detection in sequences

Under systematic perturbations on synthetic patient sequences, the model demonstrated the ability to detect different types of anomalies. It identified contextually inappropriate events, for example, cardiac medication in a trauma case reduced predictability for that event (Figure 6b). It also detected timing anomalies: premature discharge caused near-zero predictability for discharge and surrounding events, correctly flagging this as clinically impossible (Figure 6c). The model further exhibited graded temporal sensitivity, with major timing violations degrading predictability more than minor delays. Full analysis of temporal perturbations, including large delays and randomized timing, is provided in Appendix Figure A7.

**Figure 6:**
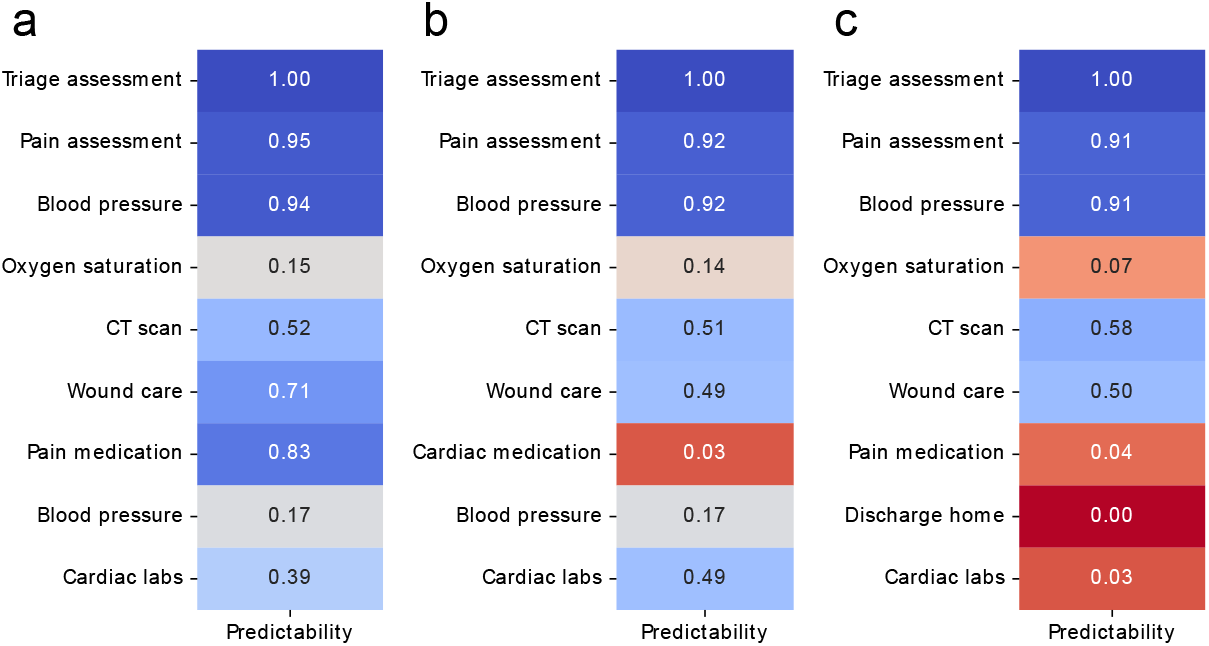
Predictability analysis of trauma pathway. (a) Standard trauma pathway showed high event predictability. (b) Inappropriate medication substitution reduced predictability. (c) Premature discharge yielded near-zero predictability scores. The model detected both context mismatches and critical pathway violations.

## 4 Discussion

Our results indicate that time-aware transformers can learn interpretable patient-specific representations from complex, irregularly timed clinical event sequences. The model simultaneously achieves strong predictive performance and enables unsupervised discovery of clinically meaningful patient subgroups in a long heterogeneous population. By bridging predictive modeling with process-oriented analysis, the framework reveals how individual events and timing patterns drive outcomes, expose pathway anomalies, and provides event-level explanations—capabilities that extend beyond those of traditional process mining and black-box deep learning approaches.

The explicit evaluation of representation quality alongside task performance has rarely been examined in prior studies [34, 35]. In our data, representations learned through self-supervision by masked event prediction captured coherent clinical patterns, yielding patient subgroups— such as high-acuity respiratory cases in younger children and complex gastrointestinal presentations in older patients — that align with clinically recognized distinctions in care pathways, resource utilization, and outcomes.

For classification, C-TAAT outperformed baselines on ESI acuity and showed competitive performance on diagnosis classification. Notably, the considerable gains in ESI acuity prediction, where temporal progression directly informs outcomes, indicate that temporal progression provides informative signals for this task, reinforcing the value of explicit time encoding. Significant improvements from self-supervised pre-training and the integration of static features further demonstrate the effectiveness of these representations across tasks.

Building on these properties, the framework enables practical applications: sequence embeddings support unsupervised subgroup discovery and resource usage analysis; event-level predictability quantifies conformance to expected care patterns; and SHAP attribution identifies which events were most influential for each prediction. Together, these analyzes extend process mining by combining pathway discovery, conformance assessment, and outcome-oriented reasoning within a single predictive framework.

In our analysis, we followed process mining conventions and restricted our data solely to event logs, excluding numerical values such as lab results or vital signs. Incorporating such values could significantly enhance predictive performance for specific outcomes and represents a promising direction for future work. Moreover, while we demonstrated the model’s capability using sequences up to 256 events, transformer architectures can inherently handle much longer sequences, suggesting that the approach could scale to more extensive patient journeys without fundamental architectural changes. Finally, while the identified subgroups closely correspond with clinically recognized care patterns, the data was primarily used to evaluate the performance and feasibility of the modeling approach. Deriving clinical insights from these findings will require further and more detailed prospective evaluation.

In conclusion, we introduced ClinicalTAAT, a time-aware representation learning framework that integrates process mining principles with deep learning to model highly variable, high-dimensional, and irregularly sampled clinical event sequences. Through self-supervised pretraining, the learned representations capture complex temporal dynamics and patient context, enabling robust downstream prediction, interpretable pattern discovery, and unsupervised identification of heterogeneous care trajectories and anomalies therein. With the focus on meaningful clinical representations, ClinicalTAAT offers a unified approach to analyzing clinical processes across settings. Time-aware transformer models can function as effective foundation models for clinical process analysis, providing a scalable basis for clinical auditing, operational optimization, and healthcare system evaluation.

## Data Availability

Sensitive patient data distribution is restricted by national and EU regulations. The authors received research approval from HUS Helsinki University Hospital (37/2023, 26/2024, 7/2025 and 12/2026). Code to generate synthetic data is available at GitHub [upon publication.]

https://github.com/ksolyom/synthetic-ehr-generator

## Author contributions

**Katalin Sólyomvári:** Methodology, Software, Investigation, Data Curation, Writing - Original Draft. **Tuomo Antikainen:** Investigation, Data Curation, Writing - Review & Editing. **Hans Moen:** Validation, Writing - Review & Editing. **Pekka Marttinen:** Validation, Writing - Review & Editing. **Risto Renkonen:** Writing - Review & Editing, Supervision, Funding acquisition. **Miika Koskinen:** Conceptualization, Methodology, Validation, Resources, Writing - Review & Editing, Project administration, Funding acquisition.

## Acknowledgements

This study was partially funded by Business Finland (grant 4911/31/2024) and received institutional funding from HUS Helsinki University Hospital (grant TYH2024242).

## Ethical considerations

Following national and EU legislation, no ethical permission was required for retrospective registry data, and the study was based on approval from HUS Helsinki University Hospital (permits HUS/§37/2023, HUS/§26/2024, HUS/§7/2025 and HUS/§12/2026). Data storage and analysis were conducted within a secure analytics platform (HUS Acamedic), ensuring patient confidentiality and compliance with the Finnish Medical Research Act for the secondary use of medical records (552/2019).

## Data availability

Sensitive patient data distribution is restricted by national and EU regulations. Code to generate synthetic data is available at GitHub [upon publication.]

## Code availability

The complete source code for ClinicalTAAT, including model implementation, pretraining pipeline, embedding extraction, and cluster visualization, is available at GitHub [upon publication] The code is released under MIT License.

