## Supplementary Material for "Learning Patient-Specific Event Sequence Representations for Clinical Process Analysis"

### Appendix A Supplementary Material

#### A.1 Synthetic data generator details

The synthetic data generator (Figure A1) produces interpretable clinical event sequences reflecting heterogeneity in patient trajectories, care intensity, and temporal dynamics observed in emergency care. The complete generation pipeline is illustrated in Figure A1.

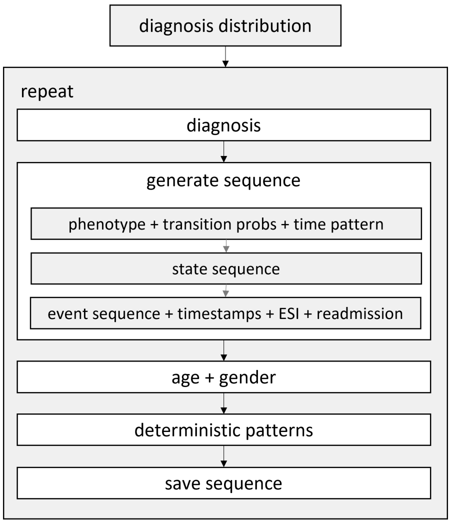


**Figure A1**: Synthetic data generation pipeline. Emergency care trajectories are simulated with distinct trajectory types, event types, and temporal dynamics using trajectory-type specific Markov processes.

The vocabulary comprises 33 event types organized into functional categories (vitals, labs, imaging, procedures, medications, consultations, discharge, critical events, administrative actions), mapped to four clinical presentations: respiratory (30%), cardiac (25%), trauma (25%), and gastrointestinal (20%).

Sequences were generated through a second-order Markov process where transition probabilities $P(e_t|e_{t-1}, e_{t-2}, \theta_p, c)$ depend on the current state, previous event, trajectory type $\theta_p$, and diagnosis category $c$. The complete trajectory-type specific transition matrices are visualized in Figure A2, illustrating the distinct clinical pathways encoded by the model.

To ensure clinical realism, the generator enforced deterministic rules: critical cardiac events trigger emergency catheterization (90% probability), and triage assessments lead to diagnosis-specific vital sign measurements. Events aligned with the patient's diagnosis with 80% probability, with diagnosis-specific profiles defining relevant event types (e.g., chest X-rays and bronchodilators for respiratory cases).

Temporal dynamics varied by care intensity: simple visits and chronic management used regular intervals drawn from an exponential distribution (mean 40 min, SD 10

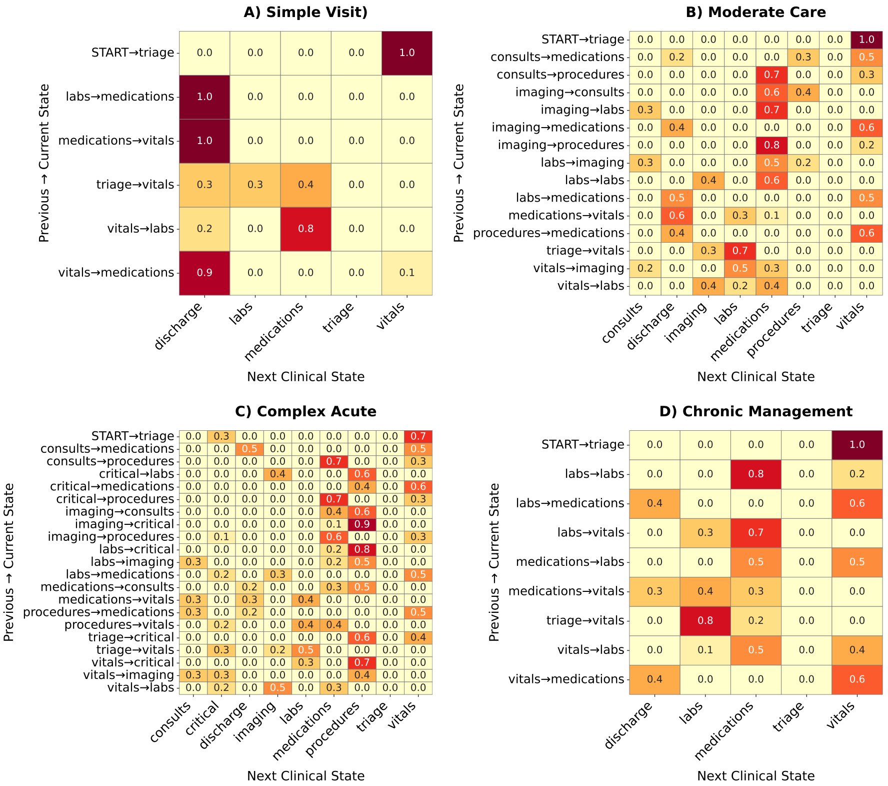


**Figure A2**: Trajectory-type specific transition matrices. Transition probabilities between clinical events differ substantially across synthetic trajectory types, confirming that the generator encodes distinct care pathways for each simulated patient group.

min). Moderate care mixed three sub-patterns: burst (exponential, mean 15 min), regular (normal, mean 40 min, SD 15 min), and sparse (randomly selecting from [0.3×, 1.0×, 2.5×] base intervals). Complex acute cases followed bimodal bursts (60% rapid interventions: exponential, mean 8 min; 40% monitoring: mean 40 min, SD 20 min). All inter-event times 2 minutes.

Each sequence included ESI scores (1–5, calculated from diagnosis acuity, trajectory type, critical events, and random variance) and diagnosis category labels (respiratory, cardiac, trauma, gastrointestinal). Static features comprised age (N(45,20), 18–90 years), gender (random binary), and 30-day readmission indicator based on diagnosis-specific profiles modulated by the severity level associated with each trajectory type. Table A1 summarizes dataset characteristics.

**Table A1**: Synthetic dataset characteristics by trajectory type.

| Property | Simple | Moderate | Complex | Chronic |
|---|---|---|---|---|
| N | 66,480 | 74,416 | 54,675 | 34,429 |
| Length | 4.6 (±0.5) | 7.4 (±2.4) | 12.9 (±5.3) | 7.4 (±2.9) |
| Span (min) | 146 (±79) | 245 (±172) | 259 (±159) | 262 (±156) |
| ESI | 3.7 (±1.2) | 2.8 (±1.2) | 1.6 (±0.8) | 2.7 (±1.2) |
| Pattern | Regular | Irregular | Burst | Regular |
| Readm. | 9.3% | 19.5% | 31.5% | 19.4% |

Continuous variables reported as mean (±SD).

#### A.2 Diagnosis category mapping

The 13 primary diagnosis categories used for prediction on real-world data were derived by grouping related ICD-10 chapters (Table A2).

**Table A2**: Mapping of ICD-10 chapters to 13 diagnosis categories used in prediction tasks.

| Diagnosis Category | ICD-10 Chapters |
|---|---|
| Infectious and parasitic diseases | A, B |
| Cancer and blood disorders | C, D |
| Endocrine, metabolic and digestive disorders | E, K |
| Mental, behavioral and nervous system disorders | F, G |
| Sensory disorders | H |
| Cardiovascular and pulmonary diseases | I, J |
| Skin and subcutaneous disorders | L |
| Musculoskeletal disorders | M |
| Genitourinary disorders | N, O, P |
| Injuries and external causes | S, T, V, W, X, Y |
| Special codes | Q, U, Z |
| Other symptoms and signs | R |
| Unclassified / other | All other chapters |

#### A.3 Training details and hyperparameters

To address significant class imbalance in both event frequencies and ESI labels, we employed focal loss [30] to down-weight frequent tokens and to focus learning on hard-to-classify instances:

$$\mathcal{L}_{\text{focal}} = -(1 - p_t)^{\gamma} \log(p_t),$$

where $p_t$ is the predicted probability of the true class and $\gamma \geq 0$ controls the focusing strength.

Hyperparameters of the pretrained model were optimized via the Optuna framework [36], using TPE sampler with early stopping (`patience`=5). The optimal model architecture (embedding dimension, number of layers, etc.) discovered during pretraining was kept fixed for finetuning. For supervised tasks, we used a reduced maximum learning rate (1e-3) to specialize the representations without overwriting the generatively learned features. We evaluated 20 configurations on a validation subset, selecting the best performing setup based on the first 15 epochs, shown in Table A3. Final training used Adam optimizer with OneCycleLR scheduler (`pct_start`=0.3), mixed precision, and gradient clipping (`max_norm`=5.0). Training was accelerated using mixed precision training via PyTorch's `Grad Scaler`, combining 16-bit and 32-bit computations for reduced memory usage and improved speed, with no performance degradation. Although the training process allowed for up to 100 epochs, early stopping was triggered at epoch 40 based on validation loss trends.

**Table A3**: Optimal hyperparameters from Optuna optimization for the pretraining task.

| **Hyperparameter** | **Value** |
|---|---|
| Embedding dimension | 64 |
| Attention heads | 2 |
| Transformer layers | 3 |
| Dropout rate | 0.23 |
| Learning rate | 0.0099 |
| Weight decay | 0.005 |
| Focal loss $\gamma$ | 1.64 |

#### A.4 Model performance on synthetic data

The pretraining phase yielded strong temporal pattern recognition (0.62 F1 masked prediction, 0.89 top-5 accuracy), demonstrating the model's ability to learn clinical workflows from unlabeled sequences. This foundation enabled accurate diagnostic classification (0.92 F1) during finetuning, confirming that diagnosis-specific patterns were well-captured in the representations. However, ESI classification remained challenging (0.36 F1), mirroring real-world difficulties in acuity assessment and suggesting that triage decisions involve complex reasoning not fully encoded in synthetic event sequences alone.

#### A.5 Comparisons with baseline models

**BEHRT for emergency care:** Following Li et al. [20], we adapted BEHRT's dual temporal embedding approach to emergency care. Since age is static in our setting, we replaced the original age buckets with discrete time intervals representing elapsed time since admission (<1 hour, 1–6 hours, 6–12 hours, up to >7 days). Inter-event intervals were similarly discretized (<10 minutes, 10–30 minutes, up to >6 hours). The architecture preserves BEHRT's multi-head attention over combined event, absolute time, interval, and positional embeddings. Contextual patient features (age, gender, readmission) were concatenated to the sequence embedding before classification.

**STraTS for emergency care:** We implemented STraTS's continuous value embedding (CVE) mechanism [25] for timestamp encoding, which projects timestamps into learnable embeddings via a two-layer feed-forward network:

$$\mathbf{e}_t = \mathrm{FFN}^t(t_i), \quad \text{where} \quad \mathrm{FFN}(x) = \mathbf{U}\tanh(\mathbf{W}x + \mathbf{b}),$$

with $t_i \in \mathbb{R}$, $\mathbf{e}_t \in \mathbb{R}^d$, and learnable parameters $\mathbf{W} \in \mathbb{R}^{d\times 1}$, $\mathbf{U} \in \mathbb{R}^{d\times d}$, $\mathbf{b} \in \mathbb{R}^d$. While the original STraTS was designed for multivariate time series with numeric values, we adapted it to categorical event sequences by omitting the value component and processing only (time, event) tuples. This preserves the model's flexible continuous-time representation while adapting it to our event-based setting. Static features were concatenated to sequence embeddings.

Hyperparameters of the baseline models were optimized identically using the Optuna framework and all models were trained using the same pretraining-finetuning protocol as C-TAAT.

##### A.5.1 Comparison with feature-based clustering baselines

To assess whether learned embeddings capture structure beyond simple feature engineering, we compared clustering of pretrained embeddings against two baselines: (1) clustering of traditional aggregated statistical features (Table A4), and (2) a random baseline where points were uniformly distributed in 2D space and clustered into the same number of groups.

**Table A4**: Statistical features used for baseline clustering comparison

| Category | Features |
|---|---|
| Static | age, gender, diagnosis |
| Statistical | sequence length, unique events, time span, time mean, time sd, event rate |
| | max event freq, mean event freq, unique event ratio, dominant event ratio |
| Frequency | event frequency vectors across event types |

Despite the rich feature set encompassing demographic, temporal, and frequency-based characteristics, statistical feature clusters showed poor separation and heavy overlap (Figure A3a). This was confirmed quantitatively by clustering metrics (silhouette: 0.37, Davies-Bouldin: 0.82), substantially worse than the 0.52 and 0.63 achieved by learned embeddings (Table A5).

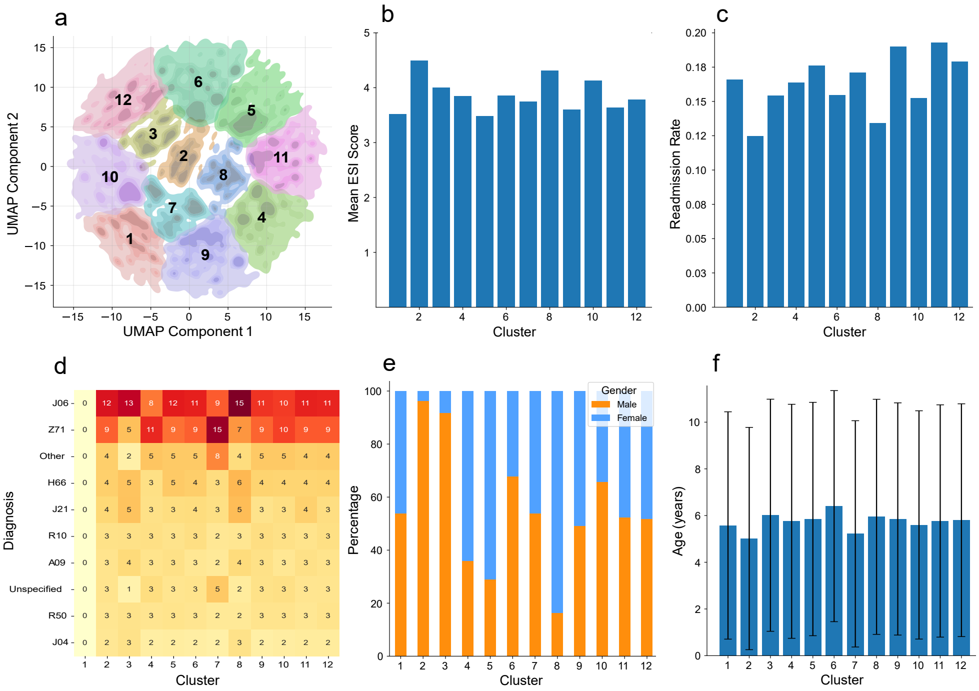


**Figure A3**: Clustering of aggregated statistical features (baseline). (a) Clustering visualization. (b) Average ESI levels per cluster. (c) Readmission rates per cluster. (d) Diagnosis diversity per cluster. Statistical features yielded less clinically coherent clusters than learned representations. (e) Gender distribution per cluster. (f) Age distribution per cluster.

**Table A5**: Clustering metrics comparison.

| **Representation type** | **Silhouette** | **Davies-Bouldin** |
|---|---|---|
| Random baseline | 0.29 | 0.80 |
| Aggregated statistical features | 0.37 | 0.82 |
| Pretrained embeddings | **0.52** | **0.63** |

All clustering is unsupervised, using pretrained-only representations.

More critically, statistical clustering failed to capture clinically meaningful variation. Clusters showed minimal differences in ESI scores, age, or diagnostic profiles (Figure A3b-f), indicating an inability to distinguish patients by severity or disease patterns. Instead, the primary distinguishing factor was gender, with extreme imbalances in some clusters. This suggests statistical features capture easily measurable attributes but fail to learn the complex relationships that characterize clinically meaningful patient subgroups. Unlike learned representations, which organize patients by medically relevant dimensions (acuity, diagnosis, resource use), statistical features produce clusters driven by demographic artifacts rather than clinically meaningful subgroups.

#### A.6 Ablation studies

##### A.6.1 Static feature integration strategies

To quantify the significance of performance differences between methods for incorporating static features, we conducted paired statistical tests using McNemar's test on the ESI classification performance. As summarized in Table A6, the cross-attention mechanism significantly outperformed both the model trained without static features ($p < 0.001$) and the simple concatenation approach ($p = 0.047$), while the difference between the model without static features and concatenation was marginally significant ($p = 0.083$). Cross-attention achieved consistent improvements across all metrics, with particularly notable gains in macro F1 (+1.6% over concatenation, +1.2% over the model without static features), suggesting better performance across all ESI acuity levels. This approach achieved 2,875 additional correctly classified cases compared to concatenation, and 3,066 compared to the model without static features. These statistically validated results demonstrate that cross-attention provides a superior framework for integrating static patient features with temporal event sequences for acuity classification.

**Table A6**: Evaluation of static feature integration strategies.

| Strategy | Accuracy | Macro F1 | Weighted F1 | p-value |
|---|---|---|---|---|
| No static info | 0.603 | 0.547 | 0.594 | |
| Concatenation | 0.607 | 0.543 | 0.596 | |
| Improvement | +0.4% | −0.4% | +0.2% | 0.08 |
| Concatenation | 0.607 | 0.543 | 0.596 | |
| Cross-attention | 0.611 | 0.559 | 0.609 | |
| Improvement | +0.4% | +1.6% | +1.2% | 0.047 |
| No static info | 0.603 | 0.547 | 0.594 | |
| Cross-attention | 0.611 | 0.559 | 0.609 | |
| Improvement | +0.8% | +1.2% | +1.5% | $< 0.001$ |

Statistical significance assessed using McNemar's test on paired predictions between pretrained and supervised-only models.

##### A.6.2 Pretraining ablation study

Self-supervised pretraining significantly improved downstream task performance compared to supervised-only training (Table A7). To assess these gains, we conducted McNemar's test on paired predictions from both models based on real-world data. For both diagnosis and ESI classification, pretraining produced consistent improvements in accuracy and macro F1. These results demonstrate that masked event prediction yields transferable representations that consistently improve supervised classification, with robust gains across tasks of varying complexity.

**Table A7**: Pretraining ablation: downstream task performance.

| Model | Accuracy | Macro F1 | p-value |
|---|---|---|---|
| **Diagnosis classification** | | | |
| Pretrain+finetune | 0.493 | 0.384 | |
| Supervised-only | 0.449 | 0.362 | |
| Improvement | +4.4% | +2.2% | $< 0.001$ |
| **ESI classification** | | | |
| Pretrain+finetune | 0.616 | 0.568 | |
| Supervised-only | 0.597 | 0.535 | |
| Improvement | +1.9% | +3.3% | $< 0.001$ |

Statistical significance assessed using McNemar's test on paired predictions between pretrained and supervised-only models.

#### A.7 Supplementary clustering results

Clustering of pretrained patient trajectory embeddings yielded distinct groups with systematic differences across multiple clinical dimensions. Cluster characteristics reflect varying clinical complexity and care intensity: average sequence lengths ranged from 4.27 to 26.98 events (Figure A4a), total encounter durations from under 2 hours to over 28 hours (Figure A4b), and readmission rates from 11% to 24% (Figure A4c)—notably, this risk stratification emerged without readmission being a training objective.

The clusters also captured demographic variation, with mean patient age ranging from 3.09 to 9.77 years across clusters (Figure A4d), while gender distribution remained balanced (48–59% male) with no extreme imbalance. Importantly, clusters mixed diverse diagnoses rather than grouping by single diagnosis types (Figure A4e), demonstrating that the embeddings capture broader clinical patterns beyond primary diagnosis codes.

Table A8 summarizes the clinical subgroups identified through unsupervised clustering of pretrained embeddings. The clusters separate naturally by acuity, with high-acuity groups (higher in ESI 1-2 cases) showing elevated readmission and resource use, and low-acuity groups (predominantly ESI 4-5 cases) representing administrative or persistent symptom encounters. Specific patterns, such as age-stratified respiratory

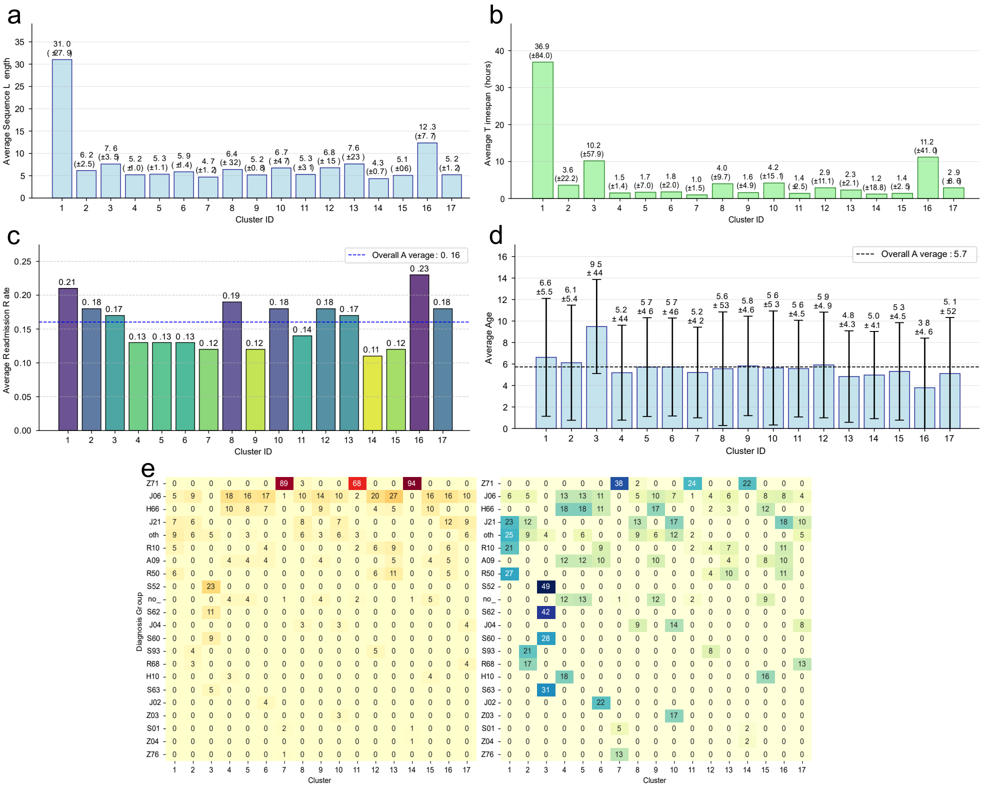


**Figure A4**: Cluster characteristics from learned embeddings. (a) Average sequence length per cluster. (b) Average episode duration per cluster. (c) Readmission rate per cluster. (d) Mean patient age per cluster. (e) Relative frequency of diagnostic categories across clusters, shown as percentages within clusters (left) and within diagnosis (right). High-acuity clusters exhibit longer sequences, longer durations, higher readmission rates, and distinct age profiles, with subgroup-specific diagnosis patterns.

infections and pediatric orthopedic trauma, demonstrate the embeddings' ability to capture clinically meaningful patient subgroups without explicit supervision.

booktabs array

**Table A8**: Characteristics of embedding-derived clusters and their associated care patterns.

| Cluster | Acuity | Age (years) | Readm. | Key Characteristics & Diagnoses |
|---|---|---|---|---|
| 1 | High | 6.6 | 21% | Older children, acute cases: abdominal pain, fever, respiratory cases. |
| 16 | High | 3.8 | 23% | Young children, acute respiratory infections, high resource use. |
| 3 | High | 9.5 | 17% | Orthopedic trauma: forearm fractures, wrist/hand injuries. |
| 2,8,10,12,17 | High | 4.2–7.1 | 15–20% | Mixed acute conditions. |
| 7,11,14 | Low | 4.9–5.8 | 11–12% | Counseling, admin (ICD-Z71), simple sequences. |
| 13 | Low | 5.1 | 17% | Persistent low-acuity symptoms: fever, gastroenteritis, URI. |
| 4,5,6,9,15 | Low | 4.5–6.2 | 13–16% | Mixed non-urgent conditions. |

Acuity: "High" = below population level average ESI (3.2), "Low" = above average ESI.
Readmission: within 30 days from discharge.

#### A.8 Supplementary experiments on synthetic data

To validate the clinical relevance of the clustering approach, we applied it to synthetic data with known trajectory types using the same pipeline as for real-world data. Analysis of the resulting clusters (Figure A5) revealed patterns consistent with our real-world findings: clusters with lower ESI scores (higher acuity) exhibited longer sequences and elevated readmission rates. For example, the highest-acuity cluster (mean ESI=1.27) had the longest care sequences, while the lowest-acuity cluster (mean ESI=4.66) captured brief, low-complexity encounters (Figure A5b–d). These results confirm that the clustering method captures clinically meaningful structure even in synthetic data, reinforcing its potential utility for identifying patient subgroups with distinct care needs and outcomes.

To quantify which events drive model predictions for different clinical tasks, we performed SHAP analysis on representative correctly-classified synthetic cases from two patient groups: cardiac and respiratory. Figure A6 shows the resulting SHAP values. For diagnosis classification, cardiac patients were most influenced by cardiac-specific events (e.g., cardiac medication), while respiratory patients were most influenced by respiratory-specific events (e.g., Airway management, Respiratory medication). For ESI classification, critical events and interventions (e.g., heart rate, intravenous access, critical cardiac event) dominated for both groups, consistent with acuity assessment prioritizing immediate actions.

To understand how different sequence modifications affect learned representations, we analyzed embedding sensitivity to controlled perturbations for ten representative synthetic patient sequences by computing Euclidean distances between original

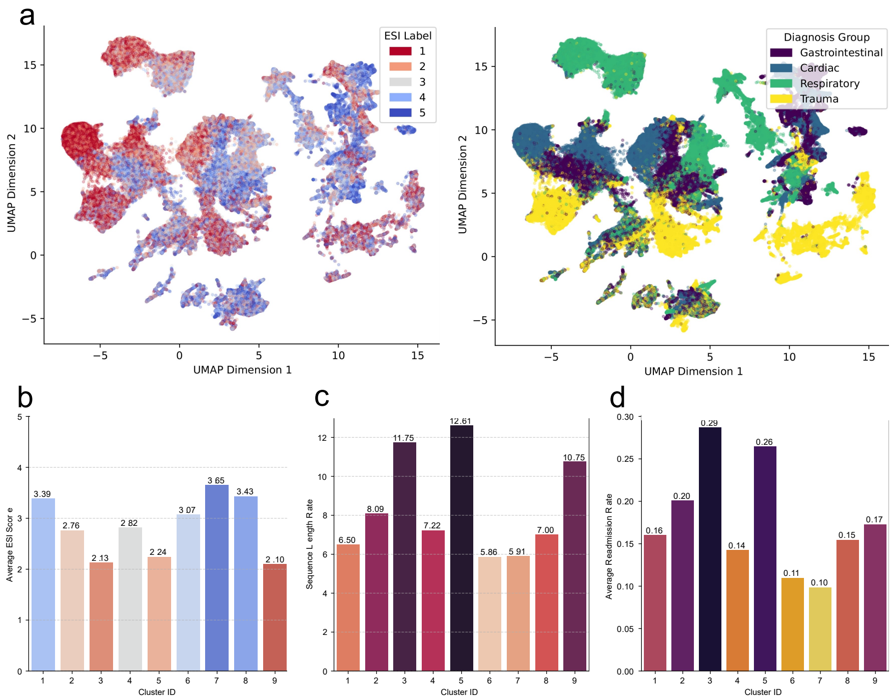

**Figure A5**: Synthetic data based cluster characteristics. (a) Embedding structure before task-specific finetuning. (b) Mean ESI score per cluster. (c) Average sequence length per cluster. (d) Readmission rate per cluster. High-acuity clusters (lower ESI) exhibit longer sequences and elevated readmission rates, confirming that the encoded synthetic care patterns are recovered by the model.

and perturbed embeddings in the reduced 2D space (Table A9). Event changes produced the largest displacements (5.3±4.0), confirming clinical content as the primary driver of representations. Other perturbations—random timing, interval increases, event shifts, and readmission changes—resulted in smaller but measurable displacements (2.7–3.0, Table A10, Figure 5). Substantial variability within each perturbation type (SD 2.2–4.0) indicates that impact depends on patient context. These findings show that while clinical content dominates, temporal modifications also influence representations, confirming the model captures both content and timing.

To further assess the model's sensitivity to timing violations beyond those shown in the main text, we analyzed two additional perturbation types on the same trauma pathway: a three-hour delay to a wound procedure and complete randomization of event timing (preserving event order) within a six-hour window. Figure A7 shows the resulting individual event predictability scores for these perturbations, which reveal that large temporal violations disrupt model confidence, demonstrating sensitivity to clinically implausible workflow timing.

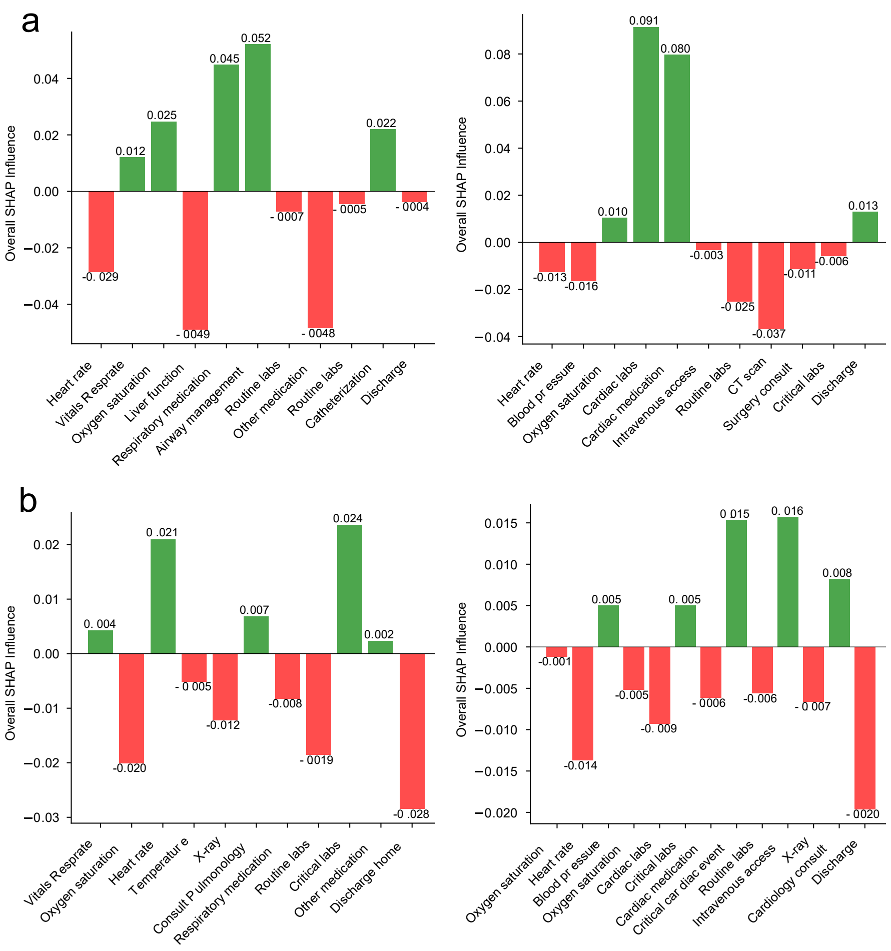


**Figure A6**: SHAP analysis of event contributions. (a) Diagnosis classification: cardiac events dominate for cardiac patients (right); respiratory-specific events dominate for respiratory patients (left). (b) ESI classification: acuity predictions are influenced by a mix of vital signs, medications, and procedures. Positive values (green) increase model confidence; negative values (red) decrease confidence. Key clinical events show strong positive influence, aligning with clinical reasoning.

**Table A9**: Types of perturbations applied to patient sequences.

| **Perturbation** | **Description** |
|---|---|
| Minor last event shift | Add 30 minutes to last event timestamp |
| Major last event shift | Add 4 hours to last event timestamp |
| Event change | Replace an event randomly with another event from the vocabulary |
| Change readmission flag | Flip readmission indicator |
| Interval increase | Add 10 minutes to all inter-event intervals |
| Random timing | Randomize event times within a 0 to 6 hours window |

**Table A10**: Embedding distances from original under sequence perturbations. Mean and standard deviation (Sd) computed across 10 representative patient cases.

| **Perturbation** | **Mean±Sd** |
|---|---|
| Event change | 5.3±4.0 |
| Random timing | 3.0±2.2 |
| Increase intervals | 2.8±2.2 |
| Major last event shift | 2.7±2.2 |
| Minor last event shift | 2.7±2.2 |
| Flip readmission flag | 2.7±2.2 |

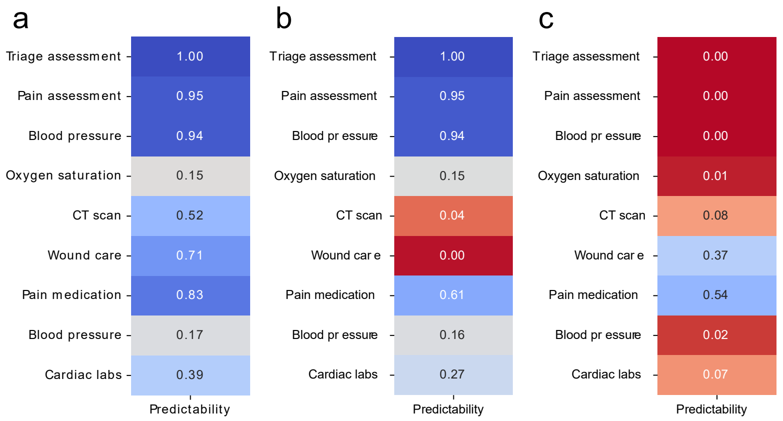


**Figure A7**: Temporal perturbation analysis on trauma pathway. (a) Original pathway shows high event predictability. (b) Three-hour delay to wound procedure reduces predictability. (c) Randomized event timing (preserving order) degrades predictability.
